# Joint modeling of survival and backwards recurrence outcomes: an analysis of factors associated with fertility treatment

**DOI:** 10.1101/2022.02.24.22271471

**Authors:** Siyuan Guo, Jiajia Zhang, Alexander C. McLain

## Abstract

The increase in methods focused on various types of survival outcomes has allowed practitioners to analyze data that are difficult or expensive to prospectively observe. Still, there are populations that are challenging to study. For example, obtaining a representative sample of couples attempting to become pregnant is difficult due to the dynamic nature of the population. This has led to an increase in the use of cross-sectional designs yielding backwards recurrent survival outcomes. In this paper, we consider the analysis of a survival outcome where subjects are observed if they are at-risk for a separate dependent survival outcome. The motivation for this problem is to determine which factors are associated with time-to-fertility-treatment (TTFT) among women currently attempting pregnancy in a cross-sectional sample. We propose appending a marginal accelerated failure time (AFT) model on TTFT with a conditional model on time-to-pregnancy (TTP) given TTFT to account for their dependence and avoid biases. We address challenges that arise due to the censoring of TTFT and the resulting increased computational complexity. The performance is validated via comprehensive simulation studies. We apply our approach to data from the National Survey of Family Growth to estimate the association insurance type has on TTFT, and estimate the impact of fertility treatment on TTP.

## 1 Introduction

Cross-sectional studies have the benefits of being low cost, time effective, and are particularly useful when studying cohorts which are difficult to sample such as dynamic populations (Vandenbroucke and Pearce, 2012). Examples of dynamic populations include couples who are attempting pregnancy (Scheike and Keiding, 2006; McLain et al., 2014), hospitalized patients after surgery (Mandel and Rinott, 2014; Mandel, 2015), and dementia patients (Bergeron et al., 2008; Carone et al., 2014). The defining feature of dynamic populations is that the length of time in the population, which we denote by *T*, is random. Commonly, it is of interest to study other outcomes within the dynamic population, which we denote by *S*. For example, we may be interested in the time-to-fertility-treatment (TTFT) among couples who are attempting pregnancy, biomarkers among hospital patients, or the time to needing in-home care among dementia patients. When *T* and *S* are dependent, this dependence needs to be accounted for to avoid biases.

There have been many different approaches proposed to analyze bivariate survival data. These include shared frailty models (Oakes, 1989; Liu et al., 2004; Hanagal et al., 2017), copula models (Genest and Rivest, 1993; Shih and Louis, 1995; Zheng and Klein, 1995; Oakes and Ritz, 2000), or semi-competing risk approaches (Fine et al., 2001; Mandel and Fluss, 2009; Mandel, 2010; McLain et al., 2021). In this paper we take an approach that is similar to the selection model framework commonly used in longitudinal analyses where the data are missing not at random (Rubin, 1976; Little and Rubin, 2019). Within this framework, the selection model consists of a marginal model for the outcome of interest and a missingness model conditional on the outcome of interest. The benefit of the selection model framework is that the model for the outcome of interest is marginal, neither the missingness pattern or a shared random effect needs to be specified for interpretation of coefficients.

Our motivating example is to study risk factors for prolonged TTFT among couples who are attempting pregnancy. Infertility is recognized as a disease of the reproductive system by the World Health Organization along with the American Society for Reproductive Medicine and is a disability under the Americans with Disabilities Act (Zegers-Hochschild et al., 2009; Practice Committee of the American Society for Reproductive Medicine, 2013). The right to procreate is considered a fundamental human right, yet large racial, ethnic, and geographic disparities persist in risk factors for infertility and for individuals who seek access to diagnostic and treatment services (Centers for Disease Control and Prevention, 2014; Farley Ordovensky Staniec and Webb, 2007). Couples attempting pregnancy is a challenging population to study since it varies over time as couples get pregnant and new couples begin their pregnancy attempts. Prospective cohort studies are difficult as they have to identify and follow couples planning to start a pregnancy attempt in the future (Scheike and Keiding, 2006; Keiding et al., 2012), and retrospective studies will sample only from people who got pregnant and thus their results suffer from lack of generalizability. Alternatively, cross-sectional studies sample a large group of individuals at high-risk for attempting pregnancy and result in a sample from the population of interest.

The benefit of using a cross-sectional study is that it results in a sample from a hard to reach group. The challenge is that the outcomes are length-biased and entirely right censored. Backwards recurrent survival methods, proposed by Cox (1969), relate the full time-to-event to the cross-sectional current duration, and been further explained in van Es et al. (2000). Keiding et al. (2002) used both parametric and nonparametric methods to estimate the survival function from current duration data. McLain et al. (2014) adjusted for time-independent covariates in the model and potential digit-preference in the outcomes. McLain et al. (2021) proposed a semi-competing risk model with a Gaussian copula of TTP and TTFT. However, their focus was on the survival function of TTP, they did not estimate the impact of the fertility treatment on the TTP or asses risk factors for prolonged TTFT.

In this paper, we propose a discrete backward recurrent survival model that incorporates time-varying covariates with time-varying coefficients. In the grouped/discrete survival framework, the time-varying coefficient model has been considered (Kauermann et al., 2005; Wong et al., 2011). Specifically, Kauermann et al. (2005) proposed a discrete survival model with time-varying coefficients and Wong et al. (2011) extended the grouped proportional hazards model with time-varying coefficient. However, there is still a gap in incorporating a time-varying covariate with a time-varying coefficient. Specifically, the length of time the time-varying covariate impacts survival may be unknown. For example, in our motivating data it is not known how long fertility treatments will impact the probability of pregnancy. As a result, we use a parsimonious form for the time-varying coefficient where the length of the impact is an estimated parameter. Further, we append this model with a marginal Accelerated Failure Time (AFT) model on TTFT (Buckley and James, 1979; Wei, 1992). This is akin to the selection model framework: a marginal model outcome of interest (here TTFT) is included with a conditional model for the missingness given the outcome of interest (here the above model for TTP given TTFT). Our main goal is to estimate risk factors for prolonged TTFT with a secondary goal of estimating the impact of the fertility treatment on TTP.

The paper is organized with the following outline. In Sections 2, we will introduce the notation, the general form of model, specify our model for the time-dependent coefficient and covariate. In Section 3, we will describe the estimation method and present results on how the computations can be simplified. We validated our model and compare it to competing approaches using comprehensive simulation studies Section 4. In Section 5, we demonstrate the proposed model on data from multiple cycles of the National Survey on Family Growth (NSFG) to determine risk factors for prolonged TTFT and the association between TTP and fertility treatment. In Section 6, we conclude with some discussion and propose future research directions.

## 2 Methods

In this Section, we develop the modeling approach that we’ll use to estimate the impact of fertility treatment on the length of pregnancy attempts. In Section 2.1, we first discuss the main concepts needed for modeling current duration data and then introduce the discrete backwards recurrent Cox model which will serve as our baseline model. In Section 2.2, we incorporate time-varying covariates with time-varying effects. In Section 2.3, we give full specification of the time-varying coefficient and baseline hazard which we use in our experiments.

### 2.1 Notation and preliminaries

As described in the introduction, the conceptual data generation is that subjects begin their at-risk period at some calendar time *σ* and are sampled at time *τ*. For those at-risk at *τ*, we observed the current duration at risk denoted by *X* = *τ* − *σ*, The goal of the estimation procedure is to provide distributional and inferential estimates for the total time at-risk denoted by *T*. For our motivating example, the goal is to estimate the distribution of TTP which is slightly different than *T*. Here, *T* corresponds to the total length of pregnancy attempt as pregnancy attempts can end for reasons other than the couple getting pregnant (e.g., giving up). It is important to note that all distributional properties are made on this *T* variable.

The difficulties in using the current duration study design are that *X* is length-biased (those with longer at-risk times are more likely to be at-risk) and entirely right-censored. The relationship between *X* and *T* is made by using two critical assumptions: (i) we assume that the probability a subject was included in the sample is proportional to their total at-risk time, and (ii) *σ* occurs according to a homogeneous Poisson process. Under these assumptions, the mass function of *X*, denoted as *g*, has form

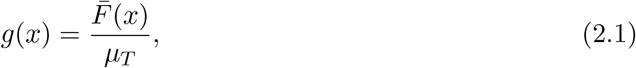

(Cox, 1969), where 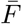 is the survival function of *T*, i.e. 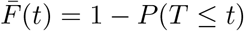, and *µ*_*T*_ = *E*(*T*) which we assume to be finite. Note that (2.1) implies *g*(0) = 1*/µ*_*T*_ and 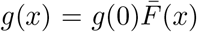. In our motivation data, the observe data are discrete so we focus throughout on this situation.

To incorporate the impact of baseline covariates ***Z*** = (*Z*_1_, *Z*_2_, …, *Z*_*P*_) on *T* we assume a discrete proportional hazards model where the hazard probability of *T* is *P* (*T* = *j*|*T* ≥ *j*) = 1 − exp[−*α*_*j*_ exp{***β***^*T*^ ***Z***}], where *α*_*j*_ ≥ 0 and *α*_0_ ≡ 0. As a result, the survivor function of *T* takes the form

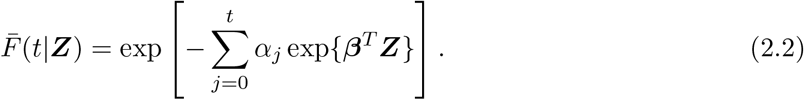

Using equation (2.1), the mass function of *X* is given by

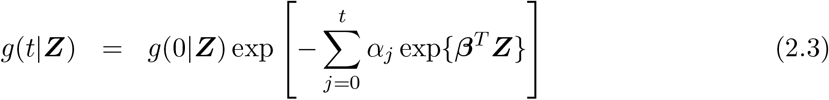

where 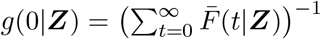. Notice *g*(*x*|***Z***) is non-increasing in *x* with maximum value at *g*(0|***Z***) and *g*^*−*1^(0|***Z***) = *E*(*T* |***Z***) is the mean of *T* given the covariate vector ***Z***. For discrete *T, β*_*j*_ can be interpreted as the approximate logarithms of subject-specific risk or odds ratios associated with a one unit increase in in *Z*_*j*_ (Ecochard and Clayton, 2000).

### 2.2 Incorporating time-varying variables and coefficients

Now we consider a random time-varying treatment which may be observed at any time point in the current at-risk period. To this end, let *S* denote the first treatment time which may be censored by *X*. The observed data is *S*\* = min(*X, S*) and *δ* = *I*(*S* ≤ *X*) where *I*(·) is the indicator function. We make the usual independent censoring assumption, which for our motivating example amounts to assuming that they survey time (*τ*) is independent of the treatment time (*S*). Further, let 𝒴_*δ*_ denote the observed treatment history given *δ*. Here, 𝒴_1_ = {***Y*** _*S*\*_, *S*\*, *δ* = 1} where ***Y*** _*S*_ = {*Y*_*j*_; *Y*_*j*_ = *I*(*j* ≥ *S*), *j* = 0, 1, …} and *Y*_*j*_ = 1 if the treatment occurred at or before time point *j* while 𝒴_0_ = {(*Y*_0_, *Y*_1_, …, *Y*_*S*\*_), *S*\*, *δ* = 0}.

We estimate the impact of the treatment on *T* while accounting for the following factors: (i) the length of the impact is unknown and to be estimated from the model, and (ii) the treatment has no impact before it occurs. That is, we allow the impact to vary over time and extend (2.3) to include this situation. More specifically, given the treatment history 𝒴_*δ*_, let *β*_*j*_(*S*) denote the effect of the time-dependent treatment at time *j* when the treatment started at time *S*. The survival function *T* given *S* is modeled by

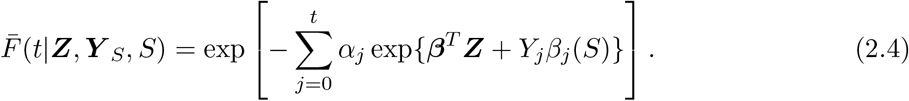

The corresponding mass function of *X* given the treatment history 𝒴_*δ*_ is

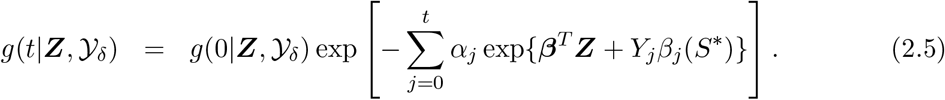

Note that we condition (2.4) on (***Y*** _*S*_, *S*) (i.e., the full treatment history) for notational convenience only, as it’s computation only requires the history up to time *t*. A more accurate representation would condition on the history up to time *t*, e.g., 𝒴_*δ*_(*t*), but this notation is more cumbersome for our purposes here. For (2.5), we condition on the observed treatment history where, if *δ*_*i*_ = 0 (2.5) can only be used for *t* = 1, …, *X*_*i*_ as the treatment information is unknown for *t > X*_*i*_ (evaluating (2.5) for *t > X*_*i*_ is not required in the likelihood calculation). We will use the notation *g*(0|***Z***, 𝒴_1_) ≡ *g*{0|***Z***, (***Y*** _*S*_, *S*)} as needed to denote (2.5) for a particular value of *S*.

As in (2.3), *g*(0|***Z***, 𝒴_*δ*_) is an expectation which requires the full treatment history and its calculation depends on *δ*. For *δ* = 1, the full treatment history is known by the monotonicity of the *y*_*j*_’s and we have 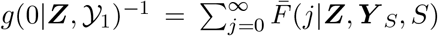. For *δ* = 0, (2.4) cannot be evaluated for *t > X* and thus

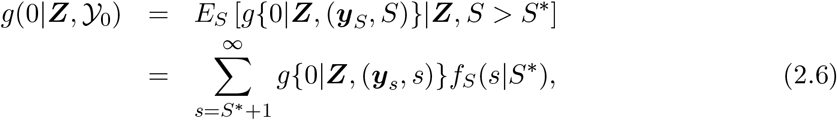

where *f*_*S*_(·|*S*\*) denotes the probability mass function of *S* given *S > S*\*. In the following Section, we discuss the specification of the distribution of *S* and adding covariates to increase the precision in the expectation in (2.6).

### 2.3 Model Specification

The above form for the model is in terms of the time-varying coefficient *β*_*j*_(·), the distribution of *S*, and the baseline hazard probability ***α***. For our motivating example, we quantify the impact of fertility treatment by assuming a parsimonious form for *β*_*j*_(·). We assume that fertility treatment has a constant impact on *T* for an estimated period of time, then has a partial impact, and then has no impact. That is, we assume the time-dependent function has the form

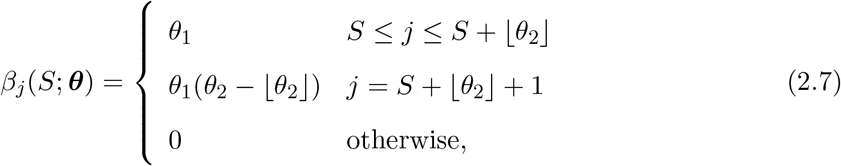

where ⌊*θ*_2_⌋ denotes the floor of the *θ*_2_ ≥ 0 and ***θ*** = {*θ*_1_, *θ*_2_}. Here, ⌊*θ*_2_⌋ + 2 represents the treatment impact length, where the first ⌊*θ*_2_⌋ + 1 time units have the full impact (*θ*_1_) followed by one time unit with a partial impact (*θ*_1_(*θ*_2_ − ⌊*θ*_2_⌋)). For *θ*_2_ = 0, the treatment effect only occurs for one time unit. While the above form is parsimonious, it does have considerable benefits and was chosen based on our motivating data. For example, it can be optimized as a continuous function of the impact *θ*_1_ and the length of the impact *θ*_2_, making it more computationally friendly than profile analytic approaches. Further, as will be discussed in Section 3, the infinite summation in (2.6) can be simplified using the properties of geometric series. Estimating the length of the treatment impact is beneficial for our motivating data as the length of time that fertility treatment has an impact is unknown.

To evaluate the likelihood the expectation in (2.6) needs to be evaluated which requires a form for the distribution of *S*. The expectation incorporates the uncertainty *S* for those where *S* is not observed. It is possible, however, to decrease the uncertainty in the distribution of *S* using available data on covariates that may be related to *S*. By incorporating covariate data into the distribution of *S* the integral in (2.6) can become more concentrated and result in less variability in the parameter estimates. By incorporating the distribution of *S* into the likelihood, the impact of the covariate data will be determined by all subjects. Let 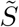 denote the continuous time to treatment variable of which we can observe 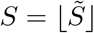. We model the distribution of 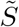 with an Accelerated Failure Time (AFT) model of time-independent covariates ***W*** = (*W*_1_, *W*_2_, …, *W*_*q*_), i.e.

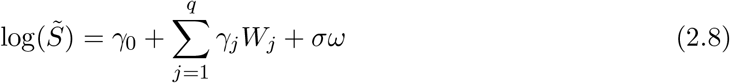

where ***γ*** = {*γ*_0_, *γ*_1_, …, *γ*_*q*_} and *ω* is the error term with a pre-specified distribution. By assuming different assumptions on the distribution of *ω*, such as standard normal, extreme value, log-gamma, the distribution of *Y* will can be changed to log-normal, Weibull and Gamma distributions, respectively. The probability mass function of *S* is then 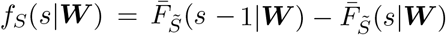 where 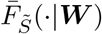 is the survival function corresponding to (2.8).

There are multiple models for ***α***, the baseline hazard probability, using either continuous or discrete hazard functions. In this paper, due to the existence of digit preference that respondents preferred to report their current duration of pregnancy attempt at months of 6, 12, 18, etc., we propose to use the piecewise constant model introduced in McLain et al. (2014). The model divides the timeline into disjoint partitions with knots ***k*** = (*k*_1_, *k*_2_, …, *k*_*m*_) and coefficients 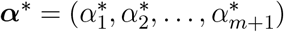 where *α*_*j*_ in (2.4) is equal 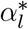 for all *j* ∈ (*k*_*l−*1_, *k*_*l*_] where *k*_0_ = 0 and *k*_*m*+1_ = ∞. Suppose *k* is a preferred digit, we will avoid knots of (*k* − 2, *k* − 1, *k, k* + 1) and with this setting, the estimation bias brought from digit preference can be weaken. Further, the knots should be chosen such that observed *S* occur in multiple intervals. Without digit preference, knots can be selected using percentiles of the observations, equal spacing between boundaries, or other approaches. AIC can be used to compare the performance of the estimation with different numbers and positions of knots.

## 3 Estimation

Using the notation of the previous section, let 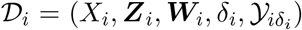 denote the observed data for subject *i* for *i* = 1, …, *n*. The parameters to be estimated are Θ = (***α, β, θ, γ***, *σ*), where ***α*** = {*α*_1_, …, *α*_*m*+1_} for a given knot set ***k***. The likelihood corresponding to the observed data is given by 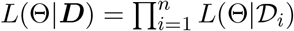 where ***D*** = {𝒟_1_, …, 𝒟_*n*_} and

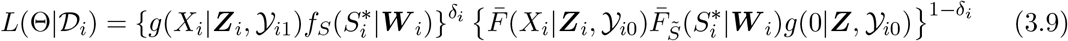

which requires the evaluation of infinite summations. For *δ* = 1, the infinite summation can be simplified by noting that under the piecewise constant model the terms in the summation of (2.4) are constant for *j > J*_*i*_ where 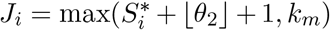. In this case, the geometric series can be used to show that

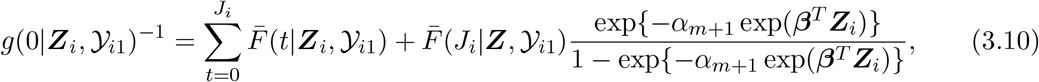

where 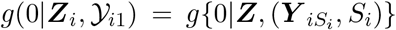. Calculating *g*(0|***Z***, 𝒴_0_) = *E*_*S*_[*g*{0|***Z***, (***y***_*S*_, *S*)}|*S > S*\*] via (2.6) requires evaluating *g*{0|***Z***, (***Y*** _*is*_, *s*)} for all *s > S*\*. This is a computationally demanding task as *J* changes with the value of *s*. A tedious but straightforward calculation can be used to show that

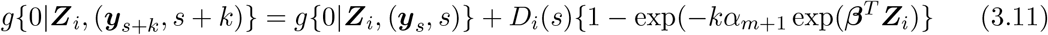

for *s > k*_*m*_ where *D*_*i*_(*s*) is a function of ***Z***_*i*_, *α*_*m*+1_, ***β*** and ***θ*** (see the supplementary material for details on this derivation). As a result, *g*{0|***Z***, (***y***_*s*_, *s*)} can be calculated via (3.10) for *s* = *S*\* + 1, …, *k*_*m*_ + 1, then (3.11) can be used for the remaining terms greatly simplifying the required calculations for (3.9). See the supplemental material for more details on how the estimation approach is implemented.

Theoretically, length of the treatment affect *θ*_2_ can be any non-negative number, but in practice setting an upper bound for *θ*_2_ is advantageous. For example, we can only realistically expect to be able to identify *θ*_2_ up to a certain point (e.g., the maximum observed survival time). In our simulations and data analyses, use a data-driven upper bound threshold 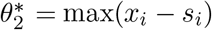, where if 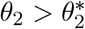 (2.4) and (2.5) are simplified by setting *β*_*j*_(*S*; ***θ***) = *θ*_1_*I*(*j* ≥ *S*).

## 4 Simulation Study

To test the performance of the proposed model and estimation procedure, numerous simulation studies were performed. The details and motivation for how the current duration *X* is obtained are given in McLain et al. (2014). Briefly, for each subject and iteration of the simulation, values of *T* were generated until their cumulative sum is greater than a large number *τ* (e.g., *τ* = 10^5^) and *X* is the difference between *τ* and the largest cumulative sum smaller than *τ* and *S* was observed if *S* ≤ *X*. Recall 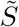 denotes the unobserved continuous time to treatment variable, and 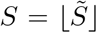 is observed data used in the model fitting. The distribution of 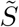 was the AFT model in (2.8), where 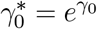, *σ*, and the distribution of *ω* were varied. Specifically, we used a log-normal *ω* with 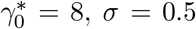 and 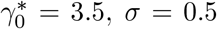, along with a Weibull *ω* with 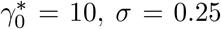 and 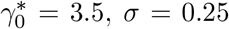. The settings with larger and smaller 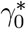 resulted in approximately 20% and 40% of *S* being observed, respectively (i.e., *E*(*δ*) ≈ 0.2 or 0.4). The AFT model of *S* and the survival function of *T* have shared two covariates, i.e, *W*_1_ = *Z*_1_ ∼Bernoulli(0.5) and *W*_2_ = *Z*_2_ ∼ *N* (0, 1). The regression coefficients are given in Table 1. The impact of *S* on *T, β*_*j*_(*S*; ***θ***), was given by (2.7) where *θ*_1_ = 1 and *θ*_2_ = 4. Given *S, T* was generated using (2.4). Sample size *n* = 1500 and 1000 iterations were used on all the simulations. To fit the proposed model, 5 and 8 knots for the log-normal and Weibull scenarios were used, respectively. The knots were chosen using the percentiles of the *X*|*δ* = 1. We compare our results to a standard AFT model on *S*, where *S* was right censored at *X* if *S > X*.

**Table 1:** Simulation results for the proposed and standard AFT models. The distribution of 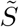 is generated from a log-normal for (a) and (b) and a Weibull for (c) and (d) where the percentage of observed *S* is approximately 20% for (a) and (c), and 40% for (b) and (d).

|  | (A) |  |  |  | (B) |  |  |  |
| --- | --- | --- | --- | --- | --- | --- | --- | --- |
|  | TRUE | MEAN | MEDIAN | SD | TRUE | MEAN | MEDIAN | SD |
| Proposed model |  |  |  |  |  |  |  |  |
| $\gamma_0^*$ | 8.0 | 8.002 | 7.98 | 0.402 | 3.5 | 3.503 | 3.499 | 0.111 |
| $\gamma_1$ | 0.5 | 0.497 | 0.498 | 0.051 | 0.5 | 0.500 | 0.499 | 0.038 |
| $\gamma_2$ | 1.0 | 0.998 | 0.998 | 0.027 | 1.0 | 1.001 | 1.00 | 0.022 |
| $\sigma$ | 0.5 | 0.498 | 0.497 | 0.020 | 0.5 | 0.500 | 0.499 | 0.015 |
| $\beta_1$ | -1.0 | -0.977 | -0.975 | 0.078 | -1.0 | -0.988 | -0.986 | 0.070 |
| $\beta_2$ | -0.5 | -0.473 | -0.473 | 0.050 | -0.5 | -0.484 | -0.483 | 0.040 |
| Standard AFT model |  |  |  |  |  |  |  |  |
| $\gamma_0^*$ | 8.0 | 9.62 | 9.61 | 0.430 | 3.5 | 4.28 | 4.27 | 0.118 |
| $\gamma_1$ | 0.5 | 0.398 | 0.398 | 0.049 | 0.5 | 0.413 | 0.413 | 0.034 |
| $\gamma_2$ | 1.0 | 0.938 | 0.938 | 0.024 | 1.0 | 0.870 | 0.870 | 0.018 |
| $\sigma$ | 0.5 | 0.480 | 0.480 | 0.019 | 0.5 | 0.452 | 0.452 | 0.013 |
|  | (C) |  |  |  | (D) |  |  |  |
| Proposed model |  |  |  |  |  |  |  |  |
| $\gamma_0^*$ | 10.0 | 9.98 | 9.98 | 0.312 | 3.5 | 3.50 | 3.50 | 0.063 |
| $\gamma_1$ | 0.5 | 0.501 | 0.502 | 0.032 | 0.5 | 0.500 | 0.501 | 0.023 |
| $\gamma_2$ | 1.0 | 1.00 | 1.00 | 0.020 | 1.0 | 0.999 | 0.999 | 0.014 |
| $\sigma$ | 0.25 | 0.249 | 0.249 | 0.011 | 0.25 | 0.249 | 0.248 | 0.009 |
| $\beta_1$ | -1.0 | -1.009 | -1.006 | 0.081 | -1.0 | -1.009 | -1.008 | 0.073 |
| $\beta_2$ | -0.25 | -0.246 | -0.245 | 0.042 | -0.25 | -0.246 | -0.248 | 0.038 |
| Standard AFT model |  |  |  |  |  |  |  |  |
| $\gamma_0^*$ | 10.0 | 11.1 | 11.1 | 0.358 | 3.5 | 4.36 | 4.36 | 0.130 |
| $\gamma_1$ | 0.5 | 0.427 | 0.428 | 0.037 | 0.5 | 0.385 | 0.385 | 0.038 |
| $\gamma_2$ | 1.0 | 0.914 | 0.916 | 0.027 | 1.0 | 0.787 | 0.789 | 0.026 |
| $\sigma$ | 0.25 | 1.27 | 1.27 | 0.016 | 0.25 | 0.277 | 0.275 | 0.022 |

Table 1 presents the mean, median, and standard deviation of regression parameters for the proposed and standard AFT models. For the proposed model, the parameters for the distribution of 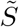 (i.e., 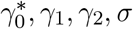) all have low bias. For the standard AFT model, there is markedly high bias in the estimates of 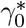 or *σ* in each setting. The percent absolute bias of the regression parameters (*γ*_1_, *γ*_2_) range from 0.0%-0.6% with the proposed approach and 6.2%-23% with the standard approach. The estimates of the regression parameters of *T* (*β*_1_, *β*_2_) were relatively unbiased.

Figure 1 shows the estimated and true baseline survival curves of *T* and *β*_*j*_(·; ***θ***) with pointwise 90% inner quantiles for the Weibull with 20% of *S* observed setting (see Section B of the Supplementary Material for other settings which were similar). Figure 1a shows the survival of *T* was estimated with little bias. The average and median values of *β*_*j*_(·; ***θ***) have low bias for *j* ≤ 4. For the remaining points the median estimate has less bias than the mean. The median estimate from *j* = 5 to 6 has a large jump, while the average estimate decreases more slowly resulting in negative bias for *j* = 5 and decreasing positive bias for *j >* 5. To have average estimates that more closely resemble the true value it appears that a larger sample size is required.

**Figure 1:**
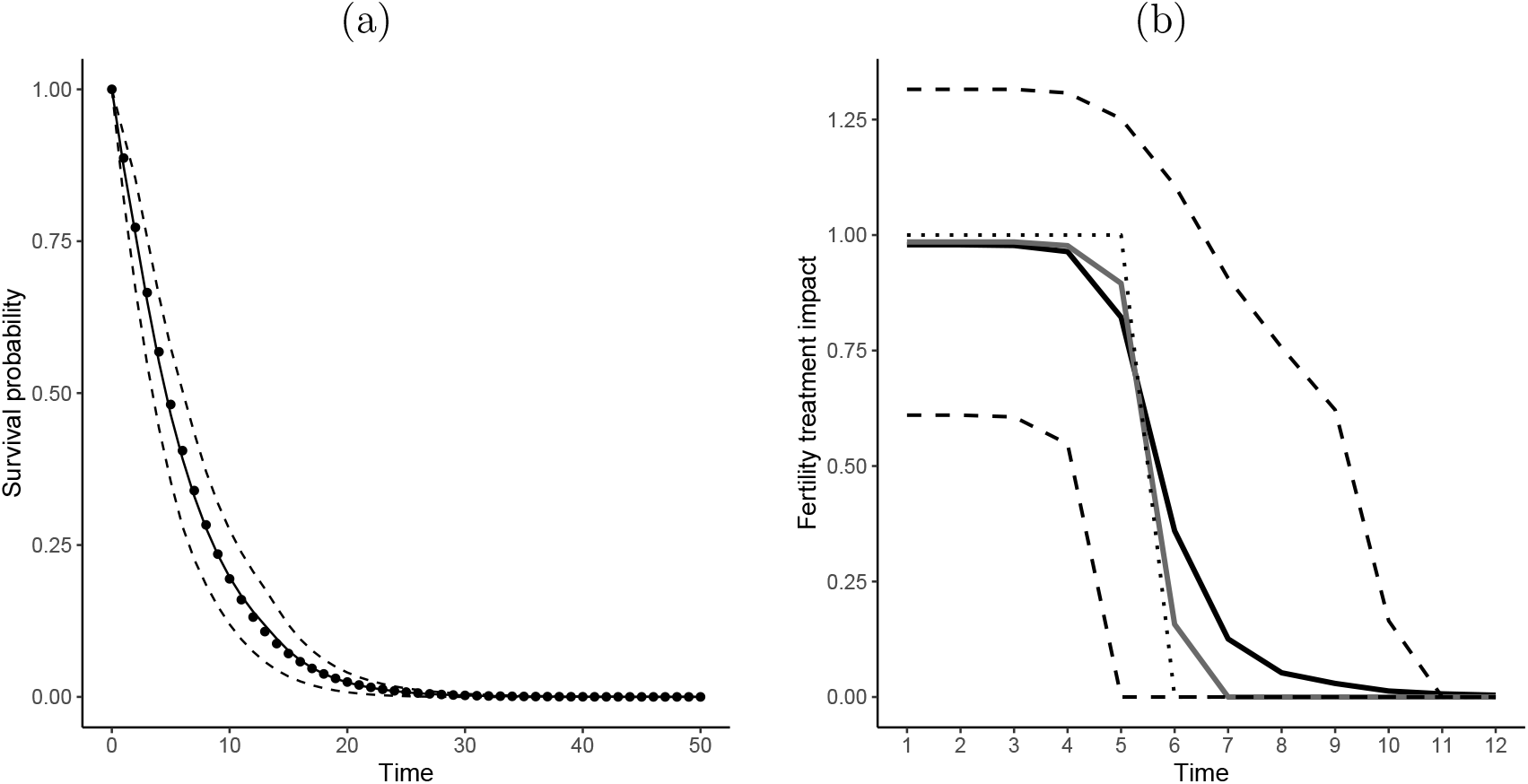
For the Weibull *S* with 40% observed setting (a) average estimated baseline survival function of *T* (solid line) with the true curves (dotted line), and (b) average (black solid) and median (gray solid) estimated *β*_*j*_(·; ***θ***) with the true value (dotted line). Both figures contain the upper and lower 5% quantiles (dashed lines).

## 5 Real data analysis

We applied the proposed model to the NSFG data which is a public survey data conducted by CDC. We integrated data from seven cycles between 2002 and 2019, which contains 1442 women that reported a current duration of pregnancy attempt. Among these women, 399 (27.6%) had undergone some type of fertility treatment. Here, we aim to determine the impact of insurance type, categorized as private (which includes military or state-sponsored health plans), medicaid/medicare, or none on TTFT. We also adjust for age (categorized *<* 35, 35 − 40, and *>* 40), and parity (yes/no). The TTP model adjusts for fertility treatment, age and parity. A log-normal assumption of *S* was chosen based on model fit criteria.

Digit preference was observed in the data such that respondents were more likely to report their current duration at the time of 6, 12, 18, 24, 36 months, etc, to minimize their impact on the estimated survival curves we chose ***k*** = (3, 8, 9, 14, 27). Further, we set 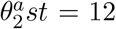 as the threshold for the impact of fertility treatment (i.e., we treat *θ*_2_ *>* 12 as a continual impact). To estimate the standard error and confidence intervals (CIs) 500 bootstrap iterations were completed. We compare our results to a standard AFT model on *S*, where if *S > X* the data were right censored at *X*.

Table 2 provides estimates and 95% CIs for the variables in the proposed and standard TTFT model. The proposed model found that the impact of parity, age 35-40, age over 40, medicaid, or no insurance lead to longer TTFT than the reference group (nulliparous, age less than 35 with private insurance). Specifically, having medicaid was associated with a time ratio (TR) of 2.39 (95% CI 1.70, 3.53) versus having private insurance while being over 40 was associated with a TR of 2.31 (95% CI 1.73, 3.25) versus being under 35. The difference in the estimated probability mass functions between the women with private insurance and medicaid is displayed in Figure 2a for nulliparous women under 35 years old, where for women with private insurance the median occurs at 19 months while for women with medicaid the median occurs at 44 months, over 2 years later.

**Table 2:** NSFG data analysis results for TTFT with the proposed and standard AFT model. Displayed is the estimated coefficients and percentile based 95% bootstrap confidence intervals.

|  | Proposed model |  |  | Standard model |  |  |
| --- | --- | --- | --- | --- | --- | --- |
|  | EST | SE | 95%CI | EST | SE | 95%CI |
| **TTFT results** |  |  |  |  |  |  |
| INTERCEPT | 3.05 | 0.10 | (2.85, 3.25) | 3.30 | 0.09 | (3.12, 3.48) |
| PARITY | 0.23 | 0.11 | (0.004, 0.43) | 0.38 | 0.11 | (0.15, 0.60) |
| AGE |  |  |  |  |  |  |
| < 35 |  |  | (REFERENT) |  |  | (REFERENT) |
| 35 – 40 | 0.33 | 0.16 | (0.001, 0.65) | 0.24 | 0.14 | (-0.04, 0.52) |
| > 40 | 0.84 | 0.17 | (0.55, 1.18) | 0.68 | 0.17 | (0.35, 1.02) |
| INSURANCE |  |  |  |  |  |  |
| PRIVATE |  |  | (REFERENT) |  |  | (REFERENT) |
| NONE | 0.72 | 0.18 | (0.37, 1.09) | 0.86 | 0.18 | (0.51, 1.22) |
| MEDICAID | 0.87 | 0.19 | (0.53, 1.26) | 1.06 | 0.20 | (0.67, 1.45) |
| $\sigma$ | 1.44 | 0.04 | (1.36, 1.52) | 1.51 | 0.05 | (1.40, 1.61) |

**Figure 2:**
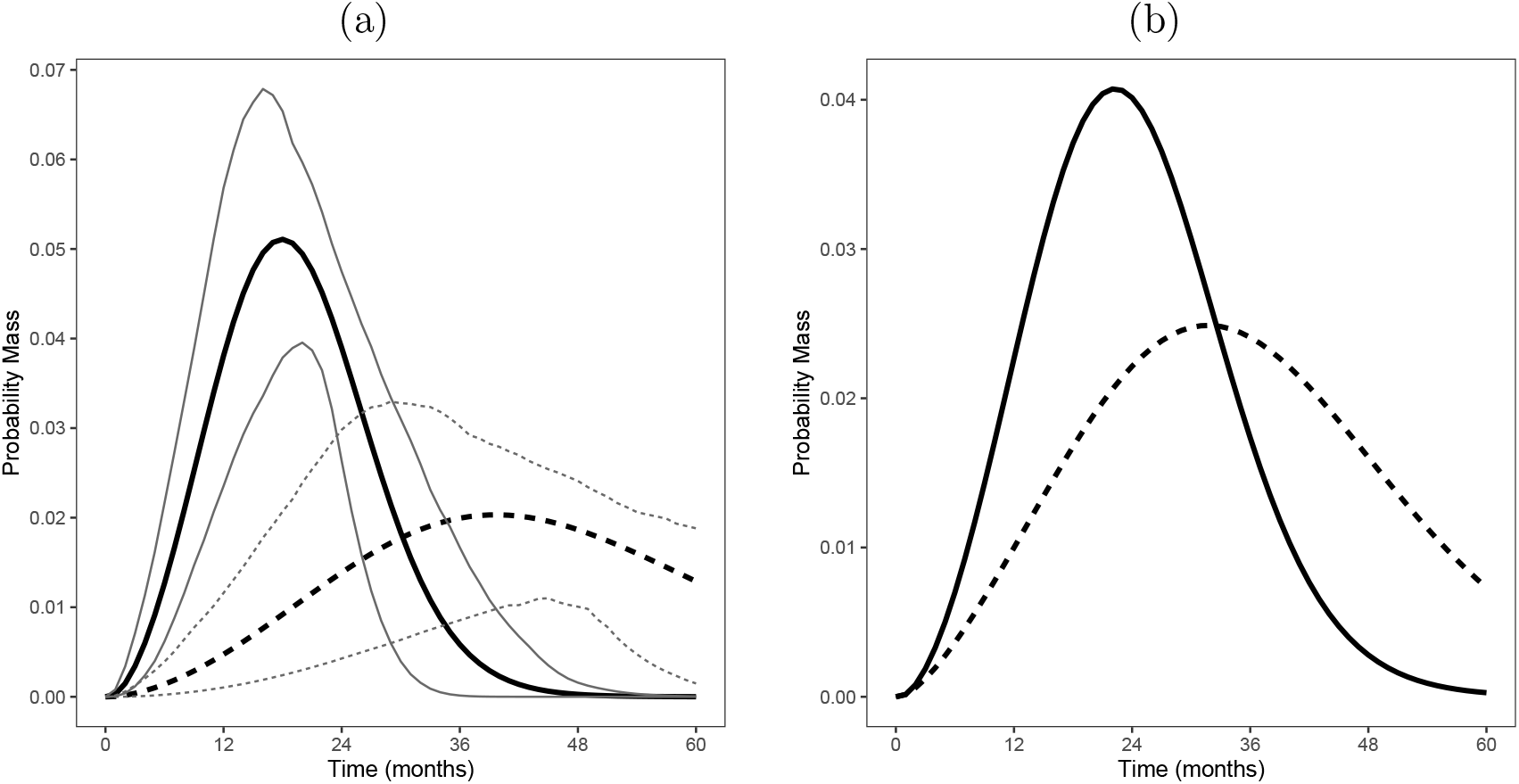
Estimated probability mass functions for TTFT (a) with 95% pointwise confidence intervals (thin dotted lines) for nulliparous women under 35 years of age with private (solid line) and medicaid (dashed line) insurance, and (b) for parous women under 35 years of age with the proposed model (solid line) and the standard AFT model (dashed line).

Generally the proposed and standard models agree on which predictor variables impact TTFT, but there are some differences in the point estimates. The impact of parity and insurance is attenuated in the proposed model versus the standard approach, while the impact of age is larger with the proposed model. For example, the TR for parity is 1.25 and 1.46 for the proposed and standard models, respectively. The estimated survival functions of TTFT for the proposed and standard model can be seen in Figure 2b for parous women under 35 years of age. For this group, the estimated median TTFT was 23 and 34 months for the proposed and standard models, respectively. We tested adding race/ethnicity to the presented model and found that it did not have an impact (data not shown).

For the TTP outcome, we found that being age 35-40 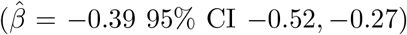 or over 40 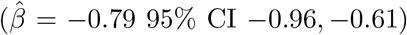 were associated with longer TTP versus under 35. Further, parous women had higher odds of getting pregnant 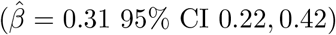 than nulliparous women. In Figure 3a, we display the estimated survival functions with 95% pointwise confidence intervals for parous women under 35 years old and nulliparous women over 40 years old (the groups with the highest and lowest odds of getting pregnant). Not including the impact of fertility treatment, the estimated median TTP for parous women under 35 years old was 10 months while for nulliparous women over 40 years old it was 28 months.

**Figure 3:**
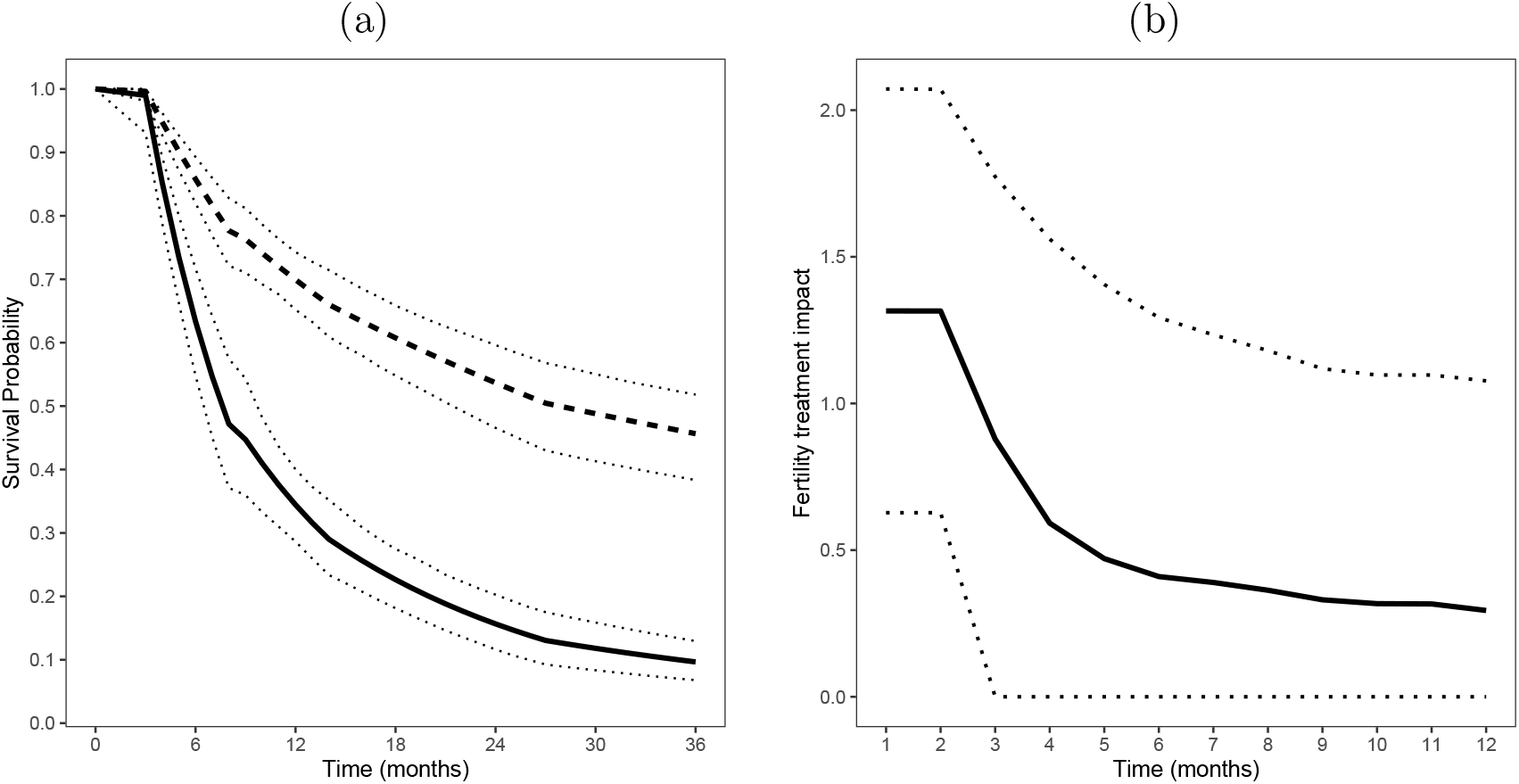
(a) Estimated survival functions for TTP with 95% pointwise confidence intervals for parous women under 35 years old (solid line) and nulliparous women over 40 years old (dashed line). (b) The estimated impact of fertility treatment *β*_*j*_(·; ***θ***) by month with 95% pointwise confidence intervals.

In Figure 3b, we show the estimated impact of fertility treatment by month with 95% pointwise confidence intervals. The estimated impact of infertility treatment is calculated as the average value of 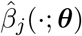 over the 500 bootstrap iterations for each *j*. The impact of fertility treatment at months 1 and 2 is clear, where the estimated coefficient is 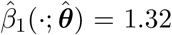 (95% CI 0.62, 2.07) for month 1 with a similar value for month 2, however, it appears to quickly diminish. For example, at month 3 the estimated coefficient is 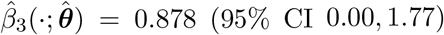 and at month 4 it is 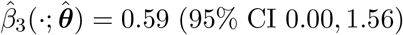. The lower bound of the CI at month 3 and after are zero. In fact, the median bootstrap 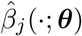 was zero for months 5 and after.

## 6 Discussion

Outcomes from dynamic populations are known to be difficult to study, arising from the random departures and entries into the set of interest. In this paper we proposed methods to analyze survival outcomes from subjects in a dynamic population, where the time in the dynamic population and the outcome of interest are dependent. We consider cross-sectional data, without follow-up, which are a cost-effective and fast method of sampling from such populations. The observed time in the population, is an example of a backwards recurrent survival outcome which require particular assumptions and methods for estimation. In this paper, we extended previous backwards recurrent methods to incorporate a time-varying covariate with a time-varying coefficient. This model was estimated jointly with an AFT model on the outcome of interest.

In the motivation data, the interest was to identify and quantify the affect of risk factors for prolonged TTFT among women attempting pregnancy and estimate the impact of fertility treatment on TTP. Risk factors associated with use of fertility treatments has been studied with similar data (see Chandra and Stephen, 2010; Greil et al., 2011; Chandra et al., 2014), however, all previous methods suffer from two major weaknesses. First, these studies use imprecise definitions of the population of interest. Chandra and Stephen (2010) and Greil et al. (2011) only consider women with fertility problems, which comes with the uncertainty of how to define “fertility problems” and how to handle couples that got pregnant quickly with fertility treatment. Further, Chandra et al. (2014) was a retrospective study of all women of reproductive age, which includes people in the that could have been in the “attempting couples” risk set decades before. Second, these studies all rely on a binary 0/1 indicator of fertility treatment. As a result, they cannot determine and quantify differences TTFT. Duron et al. (2013) used competing risks survival analysis methods to estimate the cumulative incidence of medical consultation for fecundity problems using prevalent cohort data, i.e., the subdistribution of fertility treatment and not getting pregnant. This method is different than the proposed method in the study design (prevalent cohort designs have follow-up while cross-sectional does not) and the use of competing risks methods.

In our analysis, we found that older age, being parous, and having non-private insurance lead to longer TTFT. Particularly, our model found that being over 40 years-of-age had a large impact TTFT with an TR of 2.31 (95% CI 1.73, 3.25) versus being less than 35 years-of-age. Further, having medicaid was associated with a TR of 2.39 (95% CI 1.70, 3.53) versus having private insurance. These indicate that the median TTFT for those over 40 or having medicaid was over 2.3 times larger than the referent groups. For nulliparous women under 35 years-of-age, the median TTFT occurs at 19 months with private insurance and at 44 months, over 2 years later, for women with medicaid. These types of findings were not available from previous studies that only considered a yes/no indicator of fertility treatment. After adjusting for these factors, we found that race/ethnicity did not have a significant impact on TTFT (this was also found by Chandra and Stephen, 2010). Our model also found a significant effect of fertility treatment on TTP for the first two months after treatment. However, the impact of treatment quickly diminished after month two and over half of the bootstrap estimates of the treatment impact length were less than 5 months.

While cross-sectional data are a key element to studying hard to reach populations, they do come with the drawback that the outcomes commonly come from self-recall. Such data are known to have measurement error, which has vast statistical literature. However, measurement error in the outcome is something that has garnered much less attention. Recently, Oh et al. (2021) proposed a regression calibration for this setting (see also Oh et al., 2018). In the survival setting, measurement error could increase the mean which would have an impact on the current setting since the mean is required in even the most simple models. The development of methods to correct for measurement error in current duration data is a fruitful area of future research.

## Data Availability

All data used in this study are publically available data from the National Survey of Family Growth.

https://www.cdc.gov/nchs/nsfg/

## Acknowledgements

This research was supported through the Eunice Kennedy Shriver National Institute of Child Health and Human Development grant number 1R03HD097287-01.

## Supplementary Materials for

### Section A. Likelihood function derivation details

In this Section, we’ll discuss useful forms which can be used to estimate the likelihood. Specifically, forms to estimate the likelihood for individuals with *δ*_*i*_ = 1 which is by far the more computationally demanding portion of implementing the estimation procedure. Recall, for individuals with *δ*_*i*_ = 1 the likelihood contribution is 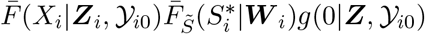, where

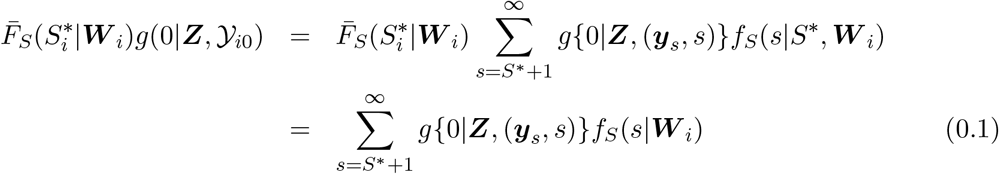

since 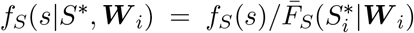. As a result, the goal is to find computationally feasible methods to calculate *g*{0|***Z***, (***y***_*s*_, *s*)} and to approximate the tail behavior of the infinite summation in (0.1).

The form for *g*{0|***Z***, (***y***_*s*+*k*_, *s*+*k*)} given in Section 3 is used when *S* is not observed and the expectation of *g*{0|***Z***, (***Y*** _*S*_, *S*)} over *S*. Here, *g*{0|***Z***, (***Y*** _*s*_, *s*)} for all *s* ≤ *k*_*m*_ must be evaluated individually. Then the form for *g*{0|***Z***, (***y***_*s*+*k*_, *s* + *k*)} is used for *s > k*_*m*_ and *k >* 0. As a result, below we assume throughout that *s > k*_*m*_. First, we define

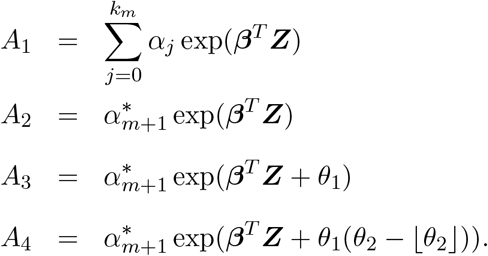

Note that when 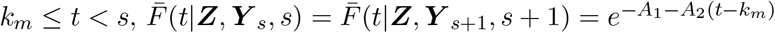. Further, if *s* ≤ *t* ≤ *s* + ⌊*θ*_2_⌋

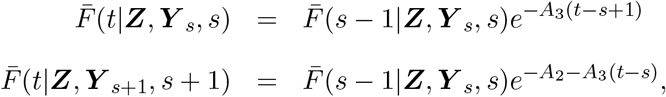

if *t* = *s* + ⌊*θ*_2_⌋ + 1,

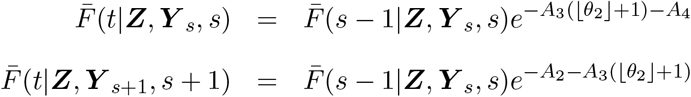

and if *t > s* + ⌊*θ*_2_⌋ + 1

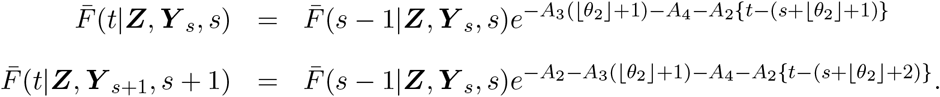

As a result, letting 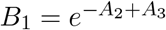 and 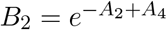, it is easy to derive

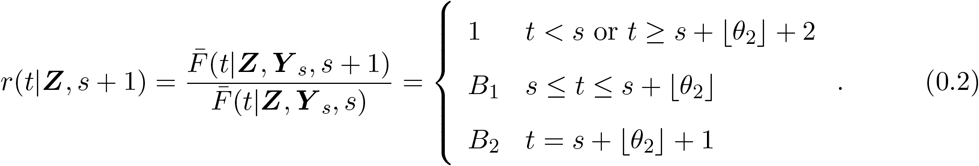

Consequently, for *s > k*_*m*_

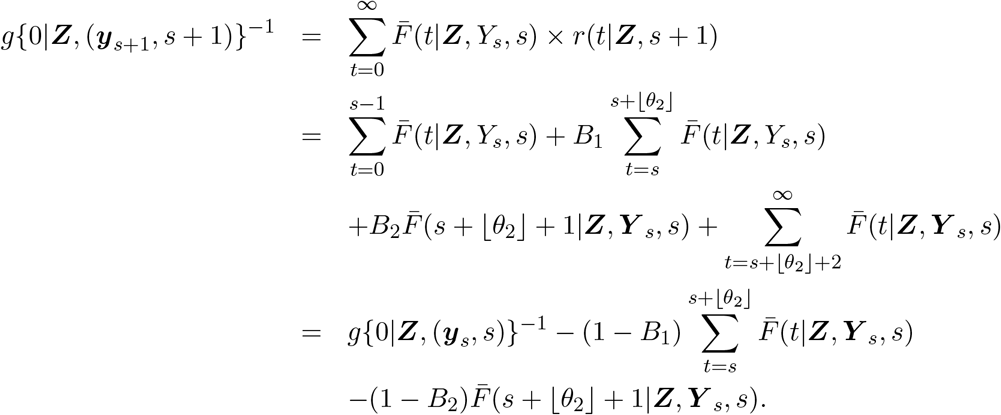

The forms for 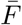 given above can be used to show that

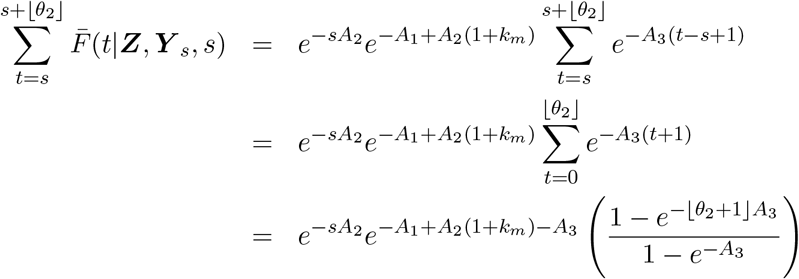

and 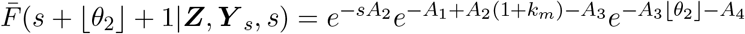. As a result,

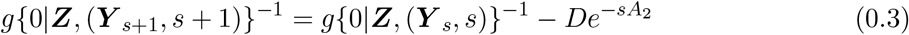

where

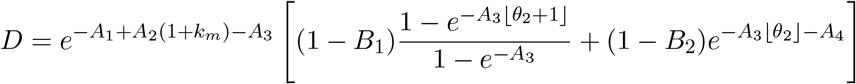

is constant in *s*. Iterating (0.3) can be used to show that

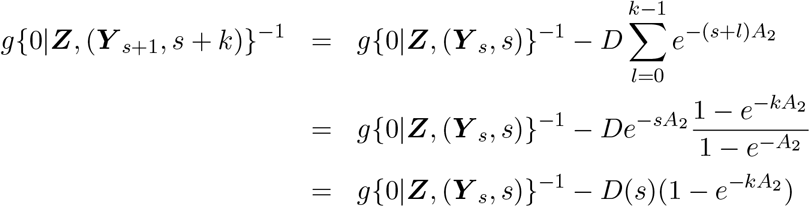

or

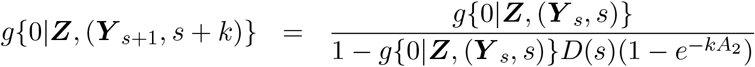

where 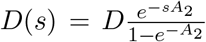, which matches the form stated in Section 3. Further, this can be used to get

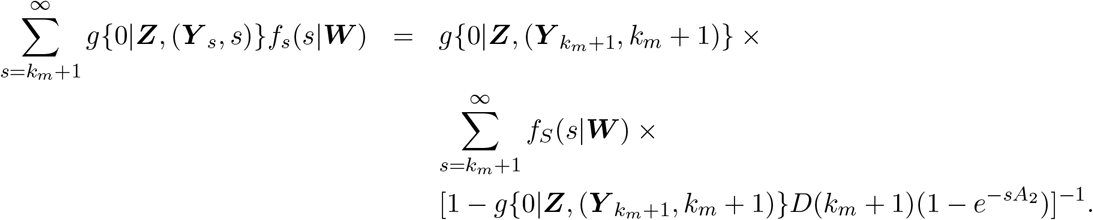

The latter summation does not have a closed form, but can be approximated by truncating it at a large number *τ*. For *S*\* *> k*_*m*_, this can be used exchanging *k*_*m*_ + 1 with *S*\* + 1. For *S*\* ≤ *k*_*m*_, the summation can be broken into [*S*\* + 1, *k*_*m*_] and [*k*_*m*_ + 1, ∞) using the above formulation. Finally, we note that when 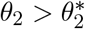,

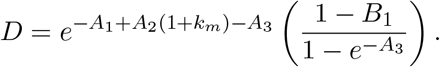

### Section B. More simulation results

In this section, we will present the remaining simulation results.

**Figure 1:**
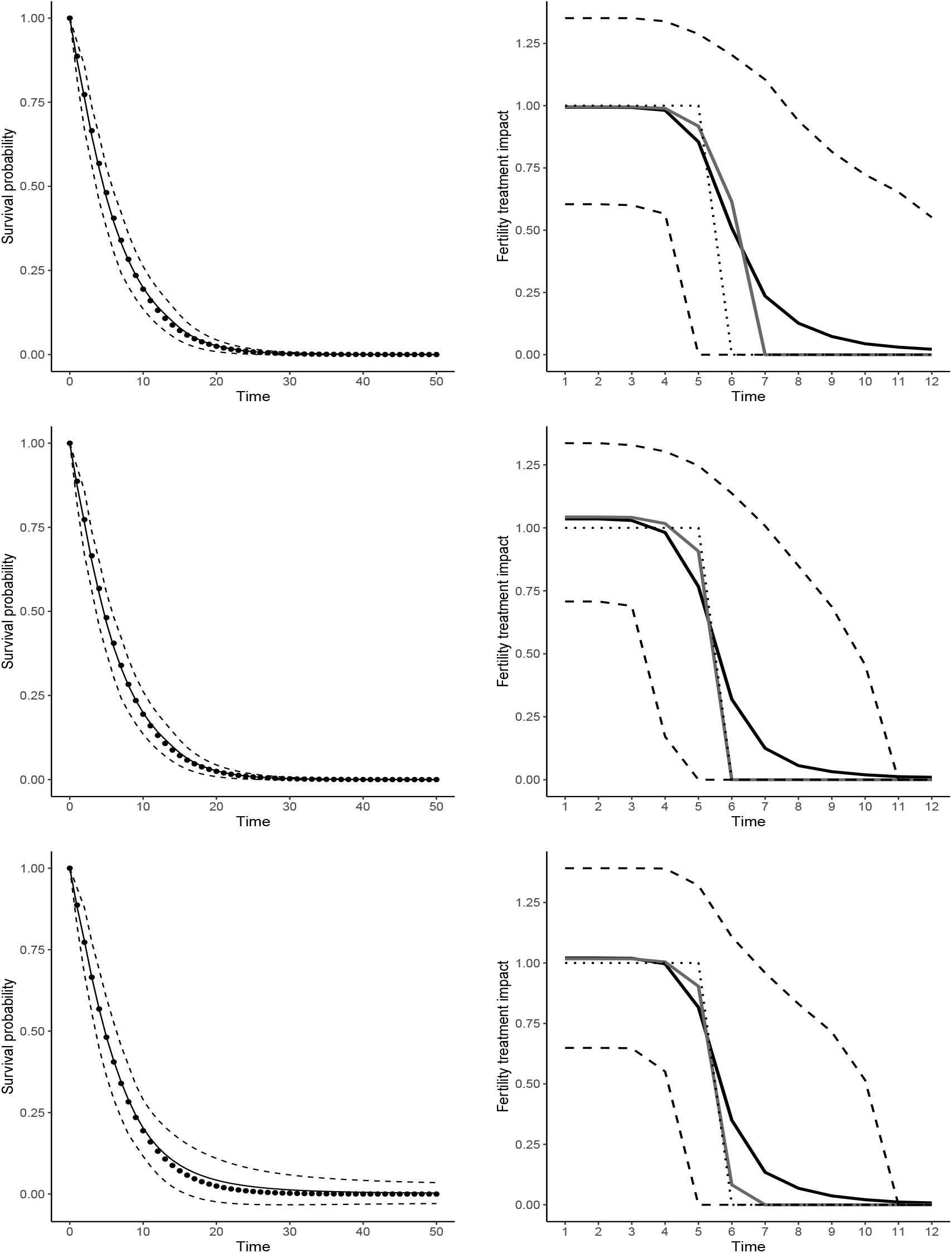
(Left) average estimated baseline survival function of *T* (solid line) with the true curves (dotted line), and (right) average (black solid) and median (gray solid) estimated *β*_*j*_(·; ***θ***) with the true value (dotted line) where all figures contain the upper and lower 5% quantiles (dashed lines). *S* is distributed as (top) log-normal with 20% observed, (middle) Weibull with 20% observed and (bottom) Weibull with 40% observed.

